# Immunogenicity and Safety of Booster Dose of S-268019-b or Tozinameran in Japanese Participants: An Interim Report of Phase 2/3, Randomized, Observer-Blinded, Noninferiority Study

**DOI:** 10.1101/2022.03.03.22271827

**Authors:** Masaharu Shinkai, Takuhiro Sonoyama, Akari Kamitani, Risa Shibata, Naomi Seki, Shinya Omoto, Masahiro Shinoda, Takashi Sato, Naoki Ishii, Kenji Igarashi, Mari Ariyasu

## Abstract

In this randomized, observer-blinded, phase 2/3 study, S-268019-b (n=101), a recombinant spike protein vaccine, was analyzed for noninferiority versus tozinameran (n=103), when given as a booster ≥6 months after 2-dose tozinameran regimen in Japanese adults without prior COVID-19 infection. Interim results showed noninferiority of S-268019-b versus tozinameran in co-primary endpoints for neutralizing antibodies on day 29: geometric mean titer (GMT) (124.97 versus 109.70; adjusted-GMT ratio [95% CI], 1.14 [0.94-1.39]; noninferiority *P*-value, <0.0001) and seroresponse rate (both 100%; noninferiority *P*-value, 0.0004). Both vaccines elicited anti-spike-protein immunoglobulin G antibodies, and produced T-cell response (n=29/group) and neutralizing antibodies against Delta and Omicron pseudovirus and live virus variants (n=24/group) in subgroups. Most participants reported low-grade reactogenicity on days 1-2, the most frequent being fatigue, fever, myalgia, and injection-site pain. No serious adverse events were reported. In conclusion, S-268019-b was safe and showed robust immunogenicity as a booster, supporting its use as COVID-19 booster vaccine.

**JRCT ID:** jRCT2031210470

**Highlights:**

- Third COVID-19 vaccine dose (booster) enhances immune response
- Interim phase 2/3 data for booster ≥6 months after the 2nd dose in Japan are shown
- S-268019-b was noninferior to tozinameran in inducing neutralizing antibodies
- Sera boosted with either vaccines neutralized Delta and Omicron virus variants
- S-268019-b was safe, and results support its use as a booster in vaccinated adults

## Introduction

Cases of coronavirus disease 2019 (COVID-19) caused by severe acute respiratory syndrome coronavirus 2 (SARS-CoV-2) are increasing periodically because of several reasons. A third vaccine dose (booster) is recommended because of concerns regarding waning humoral immunity over 6 months after the second dose [1] and consequent reduced effectiveness against SARS-CoV-2 infection [2], as well as threats from new mutant strains that may escape the vaccine-mediated immunity. Booster immunization can substantially improve the humoral immune response against the emerging variants, including Omicron [3], [4].

S-268019-b is a novel vaccine candidate comprising a modified recombinant spike protein of SARS-CoV-2 (S-910823, antigen) produced using the baculovirus expression system in insect cells and a squalene-based adjuvant (A-910823). In a double-blinded, phase 1/2 trial, S-268019-b showed tolerability and a robust immunogenicity after two doses [5]. Here, we present the interim results of a phase 2/3, randomized trial in Japan, wherein the immunogenicity and safety of a single booster dose of S-268019-b or tozinameran were assessed.

## Methods

### Study design and participants

This phase 2/3, single-center, randomized, observer-blinded, active-controlled, noninferiority trial comprised three periods: screening (day −28 to −1), evaluation (day 1 to 29), and follow-up (day 30 to 365) (**Figure 1**).

**Figure 1:**
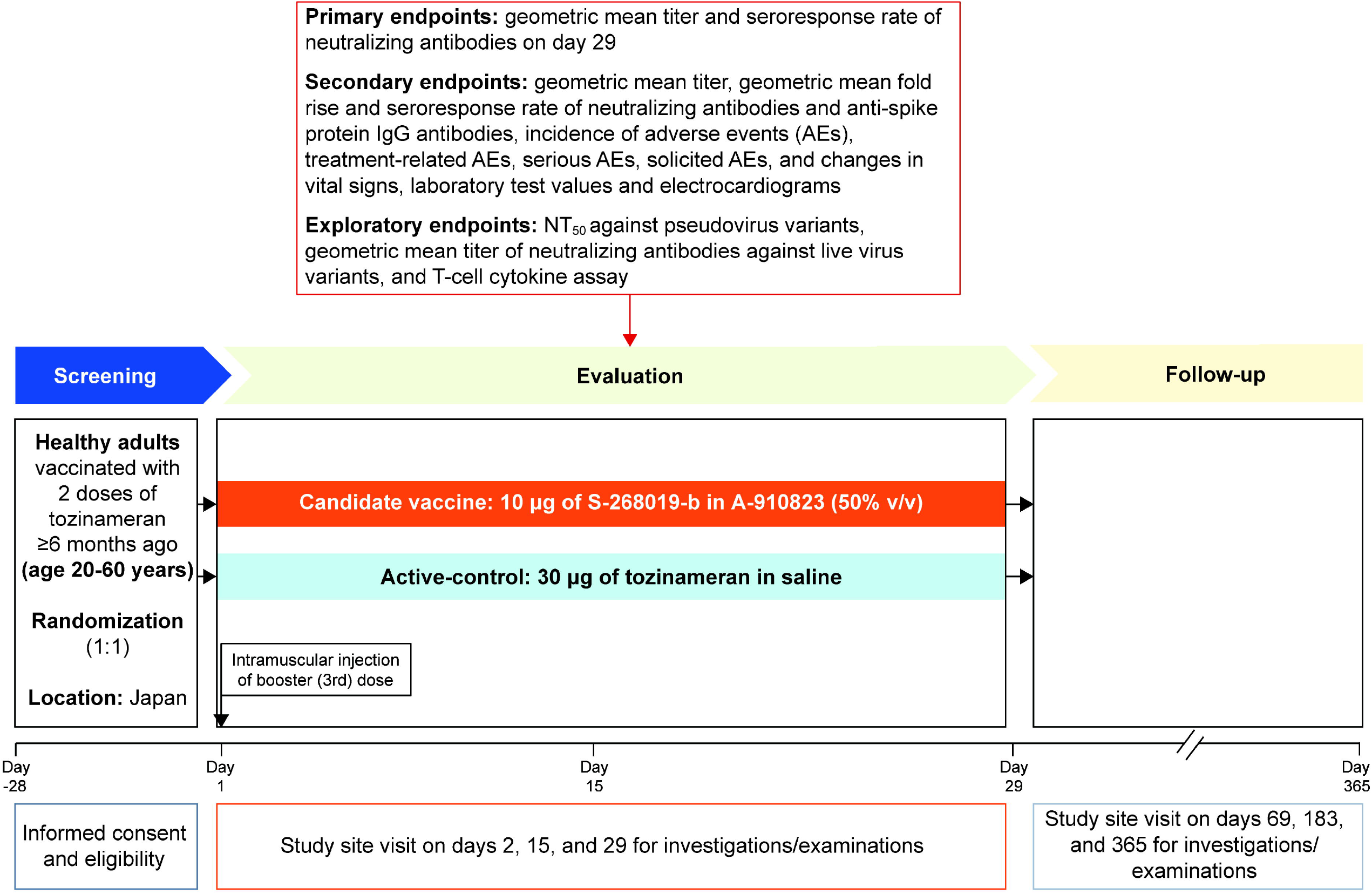
Study design, vaccine regimen, and key assessments NT_50_, 50% neutralization titer.

Participants were healthy immunocompetent Japanese adults (aged ≥20 years) who had received two doses of tozinameran (BNT162b2; *Pfizer/BioNTech* mRNA vaccine), with the second dose received ≥6 months ago. Individuals with laboratory-confirmed SARS-CoV-2 infection at screening or known history of SARS-CoV-2 infection were excluded (See **Supplementary Methods** for details).

Eligible participants were randomized 1:1, stratified by age (<40 and ≥40 years) and sex, to receive an intramuscular injection of either 0.5 mL of S-268019-b (10 μg antigen prepared with 50% v/v oil-in-water adjuvant emulsion) or 0.3 mL of tozinameran (30 μg in saline) on day 1.

The study (jRCT2031210470) was conducted in compliance with the protocol, the Declaration of Helsinki and Council for International Organizations of Medical Sciences International Ethical Guidelines, the International Council for Harmonisation of Technical Requirements for Pharmaceuticals for Human Use Good Clinical Practice Guidelines, other applicable laws and regulations, and was approved by Institutional Review Board of Tokyo Shinagawa Hospital Medical Corporation Association Tokyokyojuno-kai. All participants gave their written informed consent.

### Outcomes

The primary objective of the study was to assess the noninferiority of S-268019-b versus tozinameran as a booster dose in inducing SARS-CoV-2 neutralizing antibodies against the live wildtype virus strain (WK-521) on day 29. The co-primary endpoints included day 29 geometric mean titer (GMT) and seroresponse rate (SRR) for SARS-CoV-2 neutralizing antibodies. SRR was defined as the proportion of participants with a post-vaccination antibody titer ≥4-fold higher than the baseline.

Secondary endpoints comprised other immunogenicity parameters and safety. These included GMT, geometric mean fold rise (GMFR), and SRR for neutralizing antibodies and anti-spike protein immunoglobulin G (IgG) antibodies on days 15 and 29. Exploratory analyses in a smaller representative sample included neutralizing antibodies against SARS-CoV-2 pseudovirus variants (D614G, Delta, and Omicron) and live virus variants (wildtype, Delta, and Omicron) on day 29 and T-cell response on day 15. Safety endpoints included incidence of adverse events (AEs), serious AEs, AEs of special interest, treatment-related AEs (TRAEs), medically attended TRAEs, solicited TRAEs, and changes in laboratory test values. Immunogenicity variables with titer values below the lower limit of quantification (LLOQ) were replaced with 0.5 × LLOQ.

### Statistical analyses

The study used a noninferiority design. Noninferiority of S-268019-b to tozinameran is confirmed when the lower limit of 95% CI is >0.67 for GMT ratio (S-268019-b/tozinameran) derived from the analysis of covariate model with age and sex as covariates, and more than −10% for SRR difference (S-268019-b minus tozinameran) by the Farrington-Manning method for neutralizing antibodies on day 29 [6]. The immunogenicity subset included participants who received ≥1 dose of the study intervention, had ≥1 post-vaccination immunogenicity data, and were negative for anti-SARS-CoV-2 N-protein antibody at screening. The safety analysis subset included participants who received ≥1 dose of the study intervention. All analyses were conducted based on the actual intervention administered.

Data were summarized using measures of central tendency, dispersion, and frequency distribution. Unless otherwise noted, all statistical tests were performed at the two-sided α=0.05. Missing values were not imputed. All analyses were performed using SAS^®^ v9.4 (SAS Institute, NC, USA). (See **Supplementary Appendix** for detailed methods and statistical analyses).

## Results

### Trial Participants

All 206 participants screened were enrolled in the study during December 3– 22, 2021. Of these, two participants with unclear randomization code were excluded from the outcome analysis and 204 were analyzed (S-268019-b, n=101; tozinameran, n=103) (**Supplementary Figure 1**). Baseline demographics and participant characteristics were balanced across S-268019-b and tozinameran groups: median age (range) was 30.0 (21-59) and 31.5 (21-60) years; male population, 70% and 71%, respectively (**Supplementary Table 1**).

### Immunogenicity

GMTs (95% CIs) for neutralizing antibodies at baseline were 5.47 (4.81-6.21) for S-268019-b group and 6.65 (5.73-7.72) for tozinameran, which increased to 124.97 (108.33-144.18) and 109.70 (95.73-125.70), respectively, on day 29 (adjusted-GMTR 1.14; 95% CI 0.94-1.39; noninferiority *P-*value, <0.0001). The SRR was 100% for both groups (SRR difference 0.0; 95% CI −5.9 to 5.9; noninferiority *P-value*, 0.0004) (**Figure 2A and Table 1**). Thus, both co-primary endpoints were met: as a booster, S-268019-b was noninferior to tozinameran in SARS-CoV-2 neutralization. The GMTs (95% CIs) for anti-spike protein IgG antibodies at baseline were 1453.4 (1259.1-1677.8) for S-268019-b and 1808.2 (1546.8-2113.7) for tozinameran; these were elevated to 48464.8 (41429.9-56694.2) and 55214.8 (49013.5-62200.7), respectively, on day 29 (**Figure 2B**), with both groups showing 100% SRR (**Supplementary Table 2)**. The GMFR and GMT results were consistent (**Supplementary Table 2 and Figure 2**).

**Table 1:** Co-primary endpoints (GMT and SRR) with GMTR and SRR difference in SARS-CoV-2 neutralizing antibody response on day 29, and GMT and SRR at baseline and on day 15

| Outcome<br>(95% CI) | Tozinameran (n=102) |  |  | S-268019-b (n=101) |  |  |
| --- | --- | --- | --- | --- | --- | --- |
|  | Baseline | Day 15<sup>a</sup> | Day 29<sup>a</sup> | Baseline | Day 15 | Day 29 |
| GMT<sup>b</sup> | 6.65<br>(5.73, 7.72) | 139.48<br>(122.50, 158.82) | 109.70<br>(95.73, 125.70) | 5.47<br>(4.81, 6.21) | 127.57<br>(112.03, 145.28) | 124.97<br>(108.33, 144.18) |
| Adjusted-GMTR<sup>c</sup> | - | - | - | - | - | 1.14<br>(0.94, 1.39) |
| SRR<sup>d</sup> | - | 99.0<br>(94.6, 100.0) | 100.0<br>(96.4, 100.0) | - | 100.0<br>(96.4, 100.0) | 100.0<br>(96.4, 100.0) |
| SRR difference<sup>d</sup> | - | - | - | - | - | 0.0<br>(-5.9, 5.9) |

GMT, geometric mean titer; GMTR, geometric mean titer ratio; SARS-CoV-2, severe acute respiratory syndrome coronavirus 2; SRR, seroresponse rate.
Criteria for noninferiority confirmed when lower limit of 95% CI >0.67 for GMTR (S-268019-b/tozinameran) and > -10% for SRR difference (S-268019-b – tozinameran).
<sup>a</sup>On day 15 and day 29, tozinameran group had 101 participants. <sup>b</sup>The GMTs with corresponding 95% CIs were estimated by back transformation from the arithmetic mean and the 95% CIs based on the Student's *t* distribution of log-transformed titers to the original scale. <sup>c</sup>The adjusted-GMTR and its 95% CI were obtained using analysis of covariance model fitted on the log-transformed titers; the model included intervention group as the fixed effect as well as age (continuous) and sex as covariates. <sup>d</sup>The 95% CI were constructed using the Clopper-Pearson method for SRR and the Farrington-Manning method for SRR difference.

Furthermore, neutralizing antibodies against SARS-CoV-2 pseudovirus and live virus variants on day 29 were assessed in a representative sample selected from the immunogenicity subset (n=24/group) (**Supplementary Table 3**). Serum samples from both vaccine groups neutralized Delta and Omicron pseudovirus and live virus variants with similar potency; however, GMT against live Omicron was 4-fold lower versus wildtype (**Figure 2C and 2D**). T-cell responses were assessed for a subgroup (n=29/group) sampled from participants who gave consent for cellular immunity assessments. Both vaccines induced antigen-specific polyfunctional CD4^+^ T-cell responses, as reflected in the interferon-gamma and interleukin-2 expression on day 15 (**Supplementary Figure 2**). A strong bias toward the T-helper type 1 phenotype was noted.

**Figure 2:**
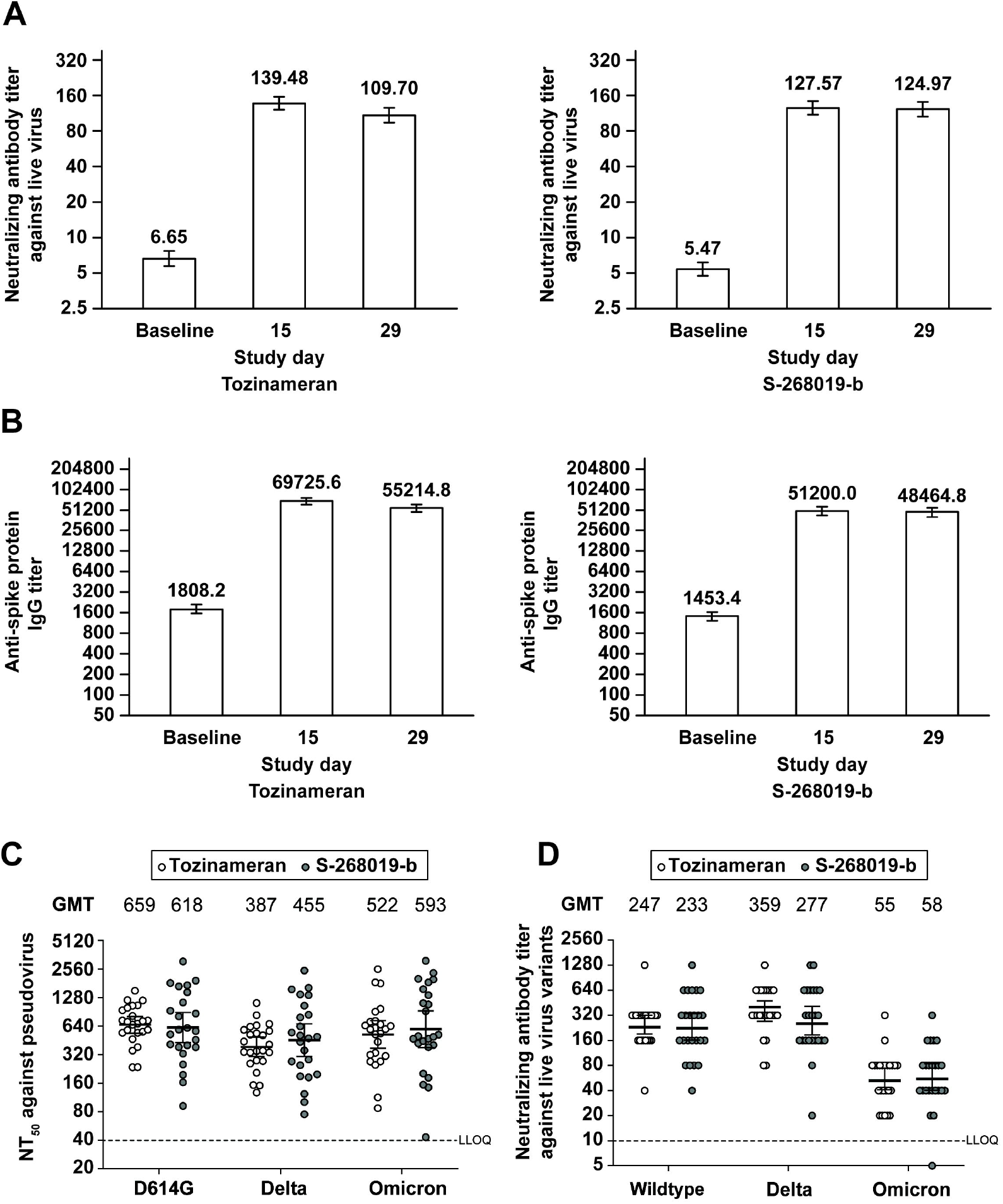
GMTs in the tozinameran and S-268019-b groups for (A) neutralizing antibodies against live wildtype virus by cytopathic effect, (B) anti-spike protein IgG, (C) NT_50_ against pseudotyped virus variants, and (D) neutralizing antibodies against live virus variants GMT, geometric mean titer; IgG, immunoglobulin G; LLOQ, lower limit of quantification; NT_50_, 50% neutralization titer. Data are presented as GMTs and 95% CIs. In Figures 2C and 2D, white and grey circles represent individual values for the tozinameran (n=24) and S-268019-b (n=24) groups, respectively, on day 29 for the same sampled cohort. The sampling ensured no significant differences in age and neutralizing antibody titer against live wildtype virus on day 29 compared with the entire cohort. Titer values reported as below the LLOQ were replaced with 0.5 × LLOQ. The LLOQ was 5 for neutralizing antibodies and 100 for anti-spike protein IgG. The 95% CI were constructed using Student’s *t* distribution for log-transformed titers.

### Safety

Both, S-268019-b and tozinameran, displayed an acceptable safety profile as a booster. There were no treatment-emergent serious AEs, deaths, grade 4-5 solicited TRAEs, or AEs of special interest reported until data cutoff date (February 4, 2022) (**Supplementary Table 4**). Overall, 96.0% (97/101) participants reported 364 TRAEs in the S-268019-b group, and 98.1% (101/103) participants reported 466 TRAEs in the tozinameran group. Furthermore, solicited systemic TRAEs were reported by 69.3% (70/101) and 79.6% (82/103) participants and solicited local TRAEs by 67.3% (68/101) and 72.8% (75/103) participants in the S-268019-b and tozinameran groups, respectively. The most frequently reported solicited TRAEs within 7 days in both booster groups were injection-site pain, fatigue, fever, myalgia, and headache (**Table 2**). Most of the solicited TRAEs were grade 1-2 and were reported on day 1-2 of the booster dose injection (**Supplementary Table 5**). One participant in the S-268019-b group and four participants in the tozinameran group experienced grade 3 solicited TRAEs.

**Table 2:** Incidence of solicited local and systemic treatment-related adverse events (experienced within 7 days after the booster) by severity in the study groups

|  | Tozinameran (n=103) |  |  |  | S-268019-b (n=101) |  |  |  |
| --- | --- | --- | --- | --- | --- | --- | --- | --- |
|  | Any grade | Grade 1 | Grade 2 | Grade 3 | Any grade | Grade 1 | Grade 2 | Grade 3 |
| Any systemic solicited TRAEs | 82 (79.6) | 47 (45.6) | 31 (30.1) | 4 (3.9) | 70 (69.3) | 55 (54.5) | 14 (13.9) | 1 (1.0) |
| Fatigue | 56 (54.4) | 33 (32.0) | 22 (21.4) | 1 (1.0) | 43 (42.6) | 34 (33.7) | 9 (8.9) | - |
| Fever | 61 (59.2) | 52 (50.5) | 7 (6.8) | 2 (1.9) | 39 (38.6) | 37 (36.6) | 1 (1.0) | 1 (1.0) |
| Myalgia | 50 (48.5) | 43 (41.7) | 7 (6.8) | - | 40 (39.6) | 39 (38.6) | 1 (1.0) | - |
| Headache | 43 (41.7) | 31 (30.1) | 12 (11.7) | - | 25 (24.8) | 19 (18.8) | 6 (5.9) | - |
| Arthralgia | 12 (11.7) | 7 (6.8) | 5 (4.9) | - | 8 (7.9) | 6 (5.9) | 2 (2.0) | - |
| Nausea/vomiting | 5 (4.9) | 5 (4.9) | - | - | 5 (5.0) | 4 (4.0) | 1 (1.0) | - |
| Diarrhea | 6 (5.8) | 4 (3.9) | 1 (1.0) | 1 (1.0) | 4 (4.0) | 3 (3.0) | 1 (1.0) | - |
| Chills | 7 (6.8) | 3 (2.9) | 4 (3.9) | - | 4 (4.0) | 2 (2.0) | 2 (2.0) | - |
| Any local solicited TRAEs (at the injection site) | 75 (72.8) | 70 (68.0) | 5 (4.9) | - | 68 (67.3) | 66 (65.3) | 2 (2.0) | - |
| Pain | 75 (72.8) | 70 (68.0) | 5 (4.9) | - | 66 (65.3) | 66 (65.3) | - | - |
| Erythema/redness | 10 (9.7) | 10 (9.7) | - | - | 6 (5.9) | 5 (5.0) | 1 (1.0) | - |
| Swelling | 1 (1.0) | 1 (1.0) | - | - | 1 (1.0) | - | 1 (1.0) | - |

TRAEs, treatment-related adverse events.
Data are presented as number (%) of participants. No participants reported grade 4 or 5 solicited TRAEs.

## Discussion

This study showed that S-268019-b as a booster (third) dose was noninferior to tozinameran in inducing SARS-CoV-2 neutralizing antibodies. Both vaccines induced neutralizing antibodies against pseudovirus and live virus variants, and elicited anti-spike protein IgG antibodies and T-cell responses within a month after the booster dose.

In other booster-dose studies, a third dose of either homologous or heterologous vaccines administered 3-9 months after the initial vaccination elicited robust immunogenicity against SARS-CoV-2 variants [7], [8], [9], [10], [11], [12]. In the COV-BOOST trial, immunogenicity of various types of vaccines given as the third dose was assessed in participants with primary vaccination with either tozinameran or AZD-1222 [12]. While the booster dose of all types of vaccines (mRNA, protein subunit, adenovirus vector, and inactivated virus) amplified immune responses, mRNA vaccines (tozinameran and mRNA-1273) induced more potent immunogenicity than other types of vaccines [12]. Considering that S-268019-b booster is as immunogenic as the tozinameran booster, S-268019-b may elicit more potent humoral immunogenicity than other types of vaccines in the COV-BOOST study, although a direct comparison is not possible between different studies.

Although no correlates of protection have been established in COVID-19, the neutralizing antibody titer against SARS-CoV-2 after primary vaccination is highly correlated with clinical efficacy against symptomatic COVID-19 [13]. Humoral immunity in COVID-19 is known to wane over time, especially after 6 months [1], and so does protection against symptomatic COVID-19 [2]. In this study, low baseline neutralizing antibody titers ≥6 months after the 2-dose tozinameran vaccination were observed, consistent with previous reports. Meanwhile, studies have reported that a booster dose of tozinameran after 2 doses of tozinameran enhances humoral immunity [7], [12]. Moreover, a report from Israel stated greater efficacy against COVID-19-related hospitalization and death after three versus two tozinameran doses [14]. Since our results show noninferiority of S-268019-b to tozinameran in inducing neutralizing antibodies, it is speculated that S-28019-b may also show clinical efficacy similar to the booster tozinameran, although further investigations to support this are warranted.

A previous study showed that the 50% neutralization titer (NT_50_) against the live Omicron virus after two doses of tozinameran was below the detection limit and 61-fold lower compared with the wildtype NT_50_ [15]. However, a booster dose of tozinameran elicited neutralizing antibody titer against Omicron, with NT_50_ only 6-fold lower than that with the wildtype [15]. Moreover, the tozinameran booster dose protected against symptomatic COVID-19 due to Omicron [16]. The current findings also indicate a 4-fold reduction in Omicron neutralizing antibodies versus wildtype; however, S-268019-b and tozinameran are similar with respect to neutralizing antibody levels against Omicron, suggesting that S-28019-b might also show similar clinical efficacy as tozinameran against symptomatic COVID-19 due to Omicron.

The T-cell response in COVID-19 is crucial for sustained protection from severe disease progression [17], [18]. Additionally, the T-cell response elicited by COVID-19 infection or prior vaccination is considered cross-reactive against variants, including Omicron [19]. In the current study, we only examined the T-cell response to the original wildtype strain, in which S-268019-b showed a response similar to tozinameran. However, due to the cross-reactive nature of the T-cell response, it is speculated that the T-cell response elicited by S-268019-b may possibly be effective in preventing severe diseases caused by Omicron and other future variants.

In this study, both vaccines led to mainly low-grade reactogenicity, with fever, fatigue, myalgia, and injection-site pain being commonly reported, usually within 2 days of the booster dose. Compared with tozinameran, S-268019-b led to a lower incidence of solicited TRAEs. Similarly, across all 7 booster vaccines in the COV-BOOST trial, the most frequently solicited AEs reported within 7 days were low-grade fatigue, headache, and local pain [12]. Despite the differences in population and methodologies across different studies, reactogenicity patterns seem largely consistent across studies after the booster dose of either an mRNA [7], [8], adenovirus vector [9], or protein subunit vaccine [10], [11], [12].

This is the first clinical trial report of a recombinant spike protein vaccine showing its noninferiority to an mRNA vaccine as a booster dose in Japanese participants. This study has a few limitations. The study (1) included only immunocompetent Japanese adults with no known history of SARS-CoV-2 infection and prior vaccination with only tozinameran, (2) had relatively small sample size, and (3) was not designed for demonstrating clinical efficacy. Also, the interim result had a short follow-up duration. Despite these limitations, a booster dose of the recombinant spike protein vaccine, S-268019-b, elicited robust immunogenicity against SARS-CoV-2, with mostly low-grade reactogenicity.

## Conclusion

The booster dose of S-268019-b vaccine was noninferior to tozinameran booster as per the findings of GMT and SRR for SARS-CoV-2 neutralizing antibodies, and was well-tolerated in fully vaccinated adult Japanese participants. S-268019-b booster was comparable with tozinameran booster in neutralizing the pseudovirus and live virus variants, Delta and Omicron. Thus, S-268019-b might be a future option for COVID-19 booster vaccine in adults.

## Data Availability

All data produced in the present study are available upon reasonable request to the authors.

## Acknowledgments

The authors thank Ms. Kiyomi Kabasawa, Ms Wakana Abe, Ms Harue Otsuka, Ms. Emi Kawabe, Ms. Narumi Iida, and Ms Etsuko Tanaka for supporting clinical trials as clinical research coordinator, and Dr. Kenichi Kamachi and all the staff of Tokyo Shinagawa Hospital for supporting the clinical trials, and Ms. Haruna Hayashi and Mr. Satoshi Kojima (Shionogi & Co., Ltd.) for supporting manuscript development. The authors also thank Dr. Minal Jaggar, PhD, Dr. Vidula Bhole, MD, MHSc, and Mr. Ivan D’Souza, MS, of MedPro Clinical Research for providing medical writing support for this manuscript, which was funded by Shionogi & Co., Ltd.

## Figure legends

**Supplementary Figure 1:**
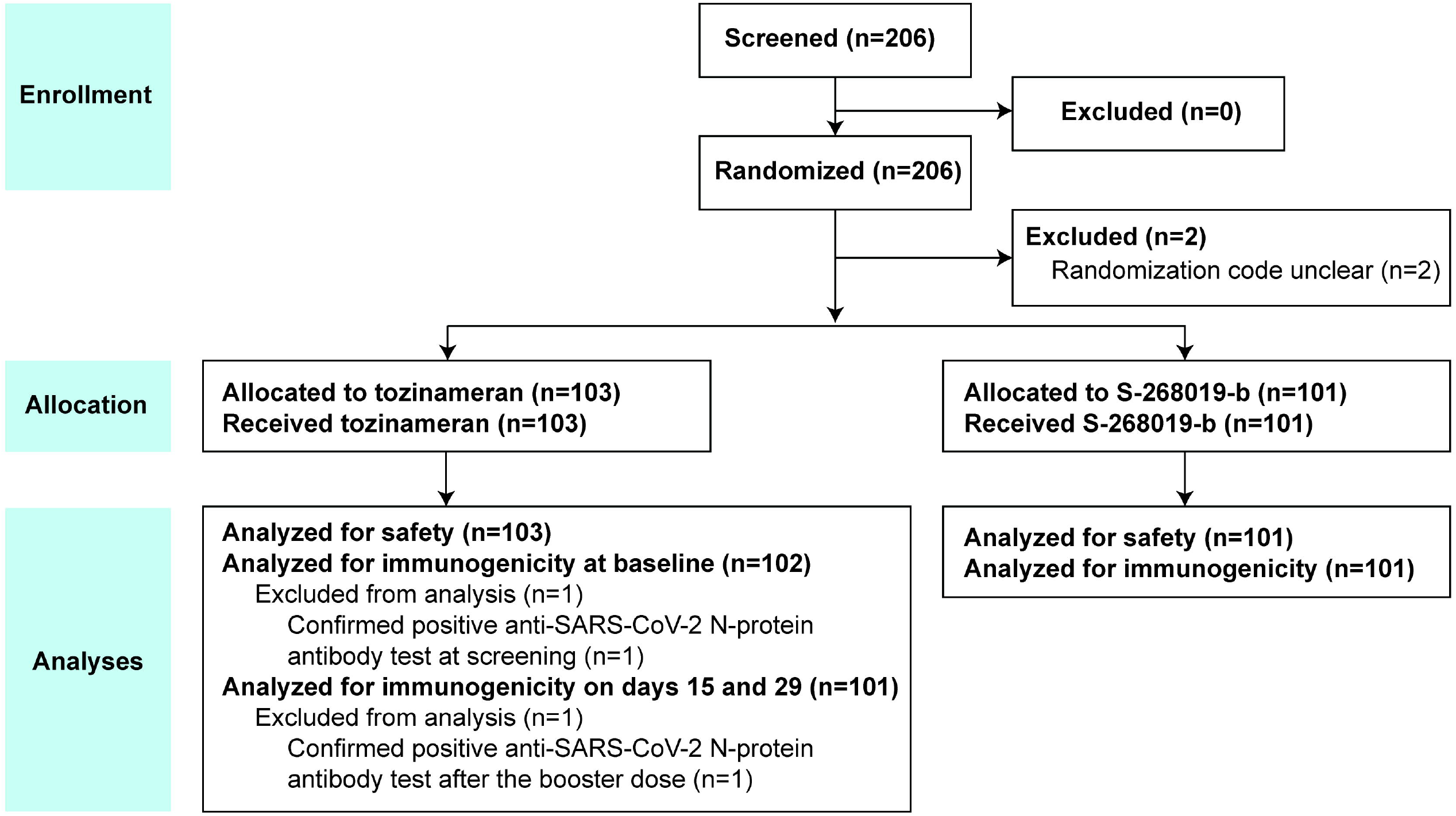
Participant flow

**Supplementary Figure 2:**
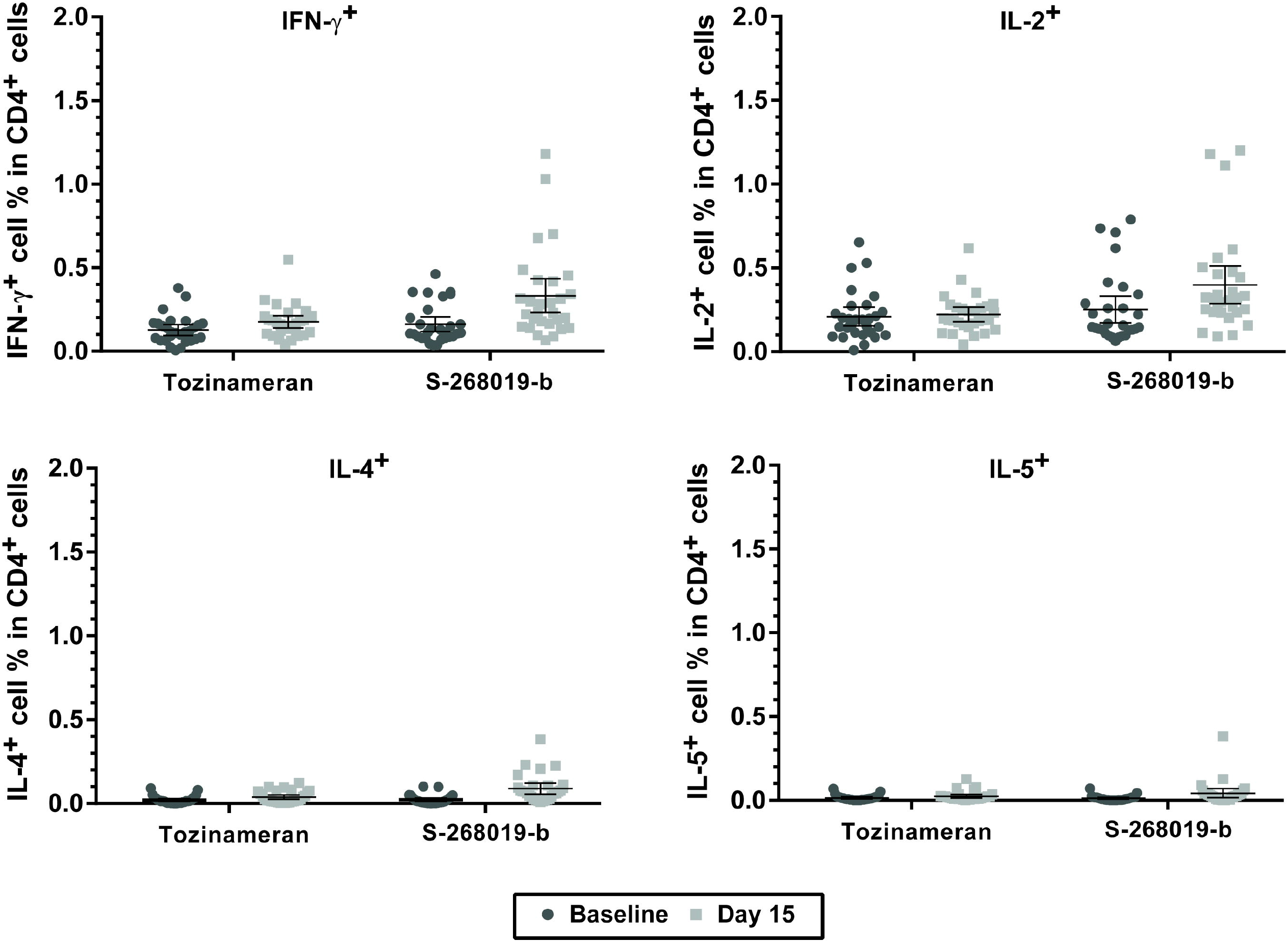
Immunologic assays from PBMCs for percent of CD4^+^ T cells positive for IFN-γ, IL-2, IL-4, and IL-5 at baseline and on day 15 in the study groups IFN, interferon; IL, interleukin; PBMCs, peripheral blood mononuclear cells. Data are presented as mean and 95% CI. Dark circles represent baseline and light squares represent day 15 values of immune marker expression for individual participants in the study groups (n=29/group). The 95% CI were constructed using the Clopper-Pearson method for proportion data.

