## Supplementary Appendix for "Immunogenicity and Safety of Booster Dose of S-268019-b or Tozinameran in Japanese Participants: An Interim Report of Phase 2/3, Randomized, Observer-Blinded, Noninferiority Study"

Supplementary Methods

*Inclusion criteria*

Participants aged 20 years or older, who had received the second dose of tozinameran ≥6 months ago, and could provide signed informed consent form were enrolled. Male participants had to refrain from donating sperm for ≥180 days after study intervention and either remain abstinent from heterosexual intercourse or use contraceptives. Female participants were eligible in the absence of pregnancy or breastfeeding and if they were not of childbearing potential or satisfied all the following conditions: used effective contraceptive method with a failure rate of <1% per year from 30 days before and ≥180 days after study intervention, agreed not to donate ova (eggs or oocytes) for the purpose of reproduction during the period from the study intervention through ≥180 days after the study intervention, and negative pregnancy test during screening.

*Exclusion criteria*

Participants were excluded from the study if any of the following criteria applied.

*Diagnostic assessments*

1. Tested positive for severe acute respiratory syndrome coronavirus 2 (SARS-CoV-2) infection as determined by SARS-CoV-2 antigen test at screening.

*Medical conditions*

2. History of SARS-CoV-2 infection, as determined in the interview before the study intervention.
3. Body temperature of 37.5°C or higher on the day of study intervention.
4. Receiving anticoagulation therapy or having thrombocytopenia or coagulopathy.
5. History of convulsion.
6. Current history of poorly controlled cardiovascular, respiratory, hepatic, renal, gastrointestinal, endocrine, hematological, or neurological disease that, in the opinion of the investigator or subinvestigator, would constitute a safety concern or confound data interpretation.
7. Immunosuppressed individuals who are immunocompromised, have acquired immunodeficiency syndrome, received steroids and systemic immunosuppressants within 6 months prior to the study intervention, are being treated for malignant tumor, or are on other immunosuppressive therapy.
8. Individuals considered having hypersensitivity to any of the study interventions or components thereof, or drug or other allergy that, in the opinion of the investigator or subinvestigator, contraindicates participation in the study, except for pollinosis and atopic dermatitis.
9. Experienced serious adverse reactions to tozinameran in the past, including myocarditis and pericarditis.
10. Having a contraindication to intramuscular injections or blood draws.

*Prior/concomitant therapy*

11. Previous SARS-CoV-2 vaccination with an approved or investigational product, except for tozinameran.
12. Any inactivated vaccine received within 14 days prior to the study intervention.
13. Any live vaccine received within 28 days prior to the study intervention.
14. Anti-SARS-CoV-2 monoclonal antibody, immunoglobulin preparations, blood products, or a blood transfusion within 3 months prior to the study intervention.

*Prior/concurrent clinical study experience*

15. Current enrollment or past participation within the last 30 days before signing of informed consent form of this study in any other clinical study involving an investigational study intervention or any other type of medical research.
16. Exposure to four or more new chemical entities within 12 months before the study intervention.

*Other exclusions*

17. Ineligibility for the study as considered by the investigator or subinvestigator.

*Cytopathic effect-based virus neutralization test*

Neutralizing antibody levels were assessed with the SARS-CoV-2 JPN/TY/WK-521 ancestral wildtype strain (kindly provided by National Institute of Infectious Diseases, Japan) and transmembrane serine protease-2 (TMPRSS-2)-expressing VeroE6 cells. Heat-inactivated vaccinated sera and convalescent plasma (56°C for approximately 30 min to remove non-specific inhibitors) were 2-fold serially diluted. Each sera/plasma sample was then mixed with 1:1 live virus suspension containing 100 times the median tissue culture infectious dose (TCID_50_)/well, and incubated for 1 h at 37°C. The mixture of sample and virus was dispensed into each well of 96-well culture plates, and VeroE6/TMPRSS2 cell suspension (1 × 10^4^ cells/well) was added to the wells. The plates were incubated at 37°C for 5 days with 5% CO_2_, followed by examination for cytopathic effect under a microscope. Virus neutralization titer was defined as the reciprocal of the highest dilution resulting in equal to or more than 50% cell viability.

*Neutralization assay against live virus variants*

Neutralizing antibody levels were assessed with the SARS-CoV-2 wildtype strain (WK-521), Delta strain (TY11-927-P1), and Omicron strain (TY38-873), kindly provided by the National Institute of Infectious Diseases, Japan, using the VeroE6/TMPRSS-2 cells. Heat-inactivated vaccinated sera and convalescent plasma (56°C for approximately 30 min to remove non-specific inhibitors) were 2-fold serially diluted. Each sera/plasma sample was then mixed with 1:1 live virus suspension containing either 2000x, 2000x, or 200,000x TCID_50_/well for wildtype, Delta, and Omicron strain, respectively, and incubated for 1 hour at room temperature for neutralization. The mixture of sample and virus was dispensed (100 µL) into each well of culture plates in duplicates, and 100 µL of VeroE6/TMPRSS2 cell suspension (3 × 10^4^ cells/well) was added to the wells. The plates were incubated at 37°C for 3 days with 5% CO_2_, followed by examination for cytopathic effect under a microscope using CellTiter-Glo^®^ 2.0 reagent (G9243, Promega) as per manufacturer’s instructions. Virus neutralization titer was defined as the reciprocal of the highest dilution resulting in equal to or more than 50% cell viability.

*Pseudotyped virus neutralization assay*

The stocks of D614G, Delta, and Omicron pseudotyped lentiviruses were diluted with the assay medium to prepare the viral suspensions of the same viral RNA copies per milliliter. Eleven serial 2-fold dilutions of heat-inactivated sera or control plasma were mixed with an equal volume of viral suspension, followed by incubating for approximately 1 h at 37°C for neutralization. After incubation, the mixture of sample and virus were added in duplicate to HEK293T stably expressing human ACE2 and TMPRSS2 cells, which were seeded into 96-well plates the day before the neutralization assay. Only virus suspension was added to Virus Control (VC) wells, which were placed on each plate. After incubating the plates at 37°C with 5% CO_2_ for 2 days, cells were lysed and subjected to luciferase assay to measure Luciferase gene expression caused by lentiviral transduction. The intensity of luminescence was measured by a microplate reader. Percent neutralization was calculated as the difference between relative light units (RLUs) of VC wells and test sample wells:

% Neutralization

= 100% × [1 − (mean RLU of duplicate sample wells ÷ mean RLU of VC wells)]

The dilution factor achieving 50% of neutralization (50% neutralization titer; NT_50_) was calculated by using the XLfit 5.3.1.3 software. When the percentage neutralization was less than 50% at the first dilution, the NT_50_ was expressed as the half of the first dilution factor. Geometric mean of the NT_50_ for each pseudovirus strain was calculated.

*Anti-spike protein immunoglobulin G (IgG) titer measurement*

Enzyme-linked immunosorbent assay (ELISA) was used to measure anti-spike protein IgG titers using duplicated samples. Full-length trimeric SARS-CoV-2 spike protein (BioServUK Ltd, BSV-COV-PR-35) was used as the immobilized antigen, and horseradish peroxidase conjugated goat-anti-human IgG (H+L) antibody (Invitrogen, A18811) was used as the detection antibody. Sample absorbance was measured as the difference between absorbance values at 405 nm and 490 nm, and the mean and coefficient of variation (CV) of absorbance for each duplicate were determined. The highest dilution factor with mean absorbance value more than or equal to the cutoff absorbance was considered the antibody titer for that sample.

*Intracellular cytokine staining (ICS) by flow cytometry*

Cytokine-producing T cells were identified by intracellular cytokine staining. Human peripheral blood mononuclear cells (PBMCs), thawed and rested for 4-5 h in R10 supplemented medium, were restimulated (1.0 × 10^6^ cells per well) with overlapping peptide pools of SARS-CoV-2 S (Miltenyi Biotec) and epitope peptide pools (Shionogi & Co., Ltd.) in the presence of Protein Transport Inhibitor Cocktail (Thermo Fisher Scientific K.K.) for 16 h at 37°C. Controls were treated with a dimethyl sulfoxide-containing medium. Cells were stained for viability and surface markers (CD3 BV421 [BioLegend]; CD4 BV510 [BioLegend]; CD8 BB515 [BD Biosciences]) in staining buffer and Brilliant Stain Buffer Plus (BSB Plus, BD Horizon, according to the manufacturer’s instructions) for 16-20 h in a refrigerator. Next, the samples were fixed and permeabilized using the Cytofix/Cytoperm kit, according to the manufacturer’s instructions (BD Biosciences). Intracellular staining for interferon gamma (IFN-γ) and interleukin (IL) (IFN-γ PE–Cy7 [BD Biosciences], IL-2 BB700 [BD Biosciences], IL-4 APC [BioLegend], and IL-5 PE [BioLegend]) was performed in the Perm/Wash buffer supplemented with BSB Plus (BD Horizon, according to the manufacturer’s instructions) for 16-20 h in a refrigerator. Samples were acquired on BD FACSCantoTM II (BD Biosciences) and analyzed with the FlowJo software version 7.6.5 (Becton, Dickinson and Company).

*Randomization*

In this study, participants whose eligibility was confirmed were randomly allocated to the S-268019-b group or the tozinameran group at 1:1 ratio by the study intervention allocation manager or the person in charge of allocation designated by the study intervention allocation manager. Participants were stratified by age (<40 years and ≥40 years) and sex. The allocation table was prepared by the permuted block randomization with a block size of 4 in each of the 4 strata. The study intervention allocation manager prepared a separate protocol for randomization and complied with it.

*Statistical Analyses*

The target sample size of 204 was determined as follows. The required number of participants to demonstrate the noninferiority of S-268019-b in geometric mean titer (GMT) of SARS-CoV-2 neutralizing antibodies and seroresponse rate (SRR) on day 29 compared with tozinameran with at least 80% power at a one-sided significance level of 0.025 is 100 participants per group, and 200 participants in total. At that time, the true geometric mean titer ratio (GMTR) between S-268019-b and tozinameran was assumed to be 1.0, and the standard deviation for the log-transformed SARS-CoV-2 neutralizing antibody titer with base 10 was assumed to be 0.4. The SRR for S‑268019-b and tozinameran were assumed to be 0.95. Considering that the Immunogenicity subset consists of participants who have a negative anti‑SARS‑CoV‑2 N-protein antibody test result at screening, and that 1 vial of tozinameran contains 6 doses, the target number of participants was set at 204.

Statistical testing for the co-primary endpoint (GMT and SRR) measurements were performed using the intersection-union test as the primary analysis. The study used a noninferiority design. The primary objective of this study was to evaluate if the immunogenicity of S-268019-b as a booster dose demonstrates noninferiority compared to tozinameran after completion of vaccination with two doses of tozinameran. The immunogenicity subset was used for the following hypothesis testing for co-primary endpoints, GMT and SRR on day 29:

Null hypothesis:	μ_A_/μ_B_ ≤ 0.67 OR r_A_ ≤ r_B_ −10%

Alternative hypothesis:	μ_A_/μ_B_ > 0.67 AND r_A_ > r_B_ −10%

where μ_A_ and μ_B_ are the geometric means of SARS-CoV-2 neutralizing antibody titers of S‑268019‑b and tozinameran on day 29, respectively, and r_A_ and r_B_ are the SRRs of SARS-CoV-2 neutralizing antibody titers of S-268019-b and tozinameran on day 29, respectively [1]. If both the lower limit of the 95% CI for the GMTR (S‑268019‑b to tozinameran) is greater than 0.67 and the lower limit of the 95% CI for the difference in SRR (S-268019-b minus tozinameran) is greater than −10%, the noninferiority is confirmed.

GMT was calculated by back transformation of the arithmetic mean of log-transformed titers. The 95% CI were constructed using Student’s *t* distribution for log-transformed titers and the Clopper-Pearson method for proportion data. For specific antibody titer at a given timepoint, the GMTR was estimated by back transformation of the intervention difference (S-268019-b minus tozinameran) and its 95% CI obtained using the analysis of covariance (ANCOVA) model fitted on log-transformed titers. The model included intervention group as fixed effect, and age (continuous) and sex as covariates. The difference in SRR between intervention groups and the corresponding 95% CI was estimated using the Farrington-Manning method.

Geometric mean fold rise (GMFR) was calculated by back transformation of the arithmetic mean of the change from baseline in log-transformed titers, where

change from baseline = (log titer at each time point) – (log titer at baseline).

**Supplementary Figure Legends**

**Supplementary Figure 1:** Participant flow

**Supplementary Figure 2:** Immunologic assays from PBMCs for percent of CD4^+^ T cells positive for IFN-γ, IL-2, IL-4, and IL-5 at baseline and on day 15 in the study groups

IFN, interferon; IL, interleukin; PBMCs, peripheral blood mononuclear cells.

Data are presented as mean and 95% CI. Dark circles represent baseline and light squares represent day 15 values of immune marker expression for individual participants in the study groups (n=29/group). The 95% CI were constructed using the Clopper-Pearson method for proportion data.

**Supplementary Table 1:** Baseline demographics and characteristics of the trial population by study groups

| **Immunogenicity subset^a^** | **Tozinameran**  **n=102** | **S-268019-b**  **n=101** |
| --- | --- | --- |
| Age (years) |  |  |
| Median (range) | 31.5 (21-60) | 30.0 (21-59) |
| <40 years, n (%) | 79 (77.5) | 77 (76.2) |
| ≥40 years, n (%) | 23 (22.5) | 24 (23.8) |
| Sex: male, n (%) | 72 (70.6) | 71 (70.3) |
| Race, n (%) |  |  |
| Asian | 102 (100.0) | 101 (100.0) |
| Ethnicity, n (%) |  |  |
| Not Hispanic or Latino | 102 (100.0) | 101 (100.0) |
| Body mass index (kg/m^2^), mean (SD) | 23.07 (4.73) | 23.24 (4.55) |
| Previous SARS-CoV-2 infection, n (%) | 0 | 0 |
| Previous SARS-CoV-2 vaccination, n (%) | 102 (100) | 101 (100) |
| Time since second dose of SARS-CoV-2 vaccine of primary series, n (%) ^b^ |  |  |
| ≥6 to <7 months | 44 (43.1) | 50 (49.5) |
| ≥7 to <8 months | 57 (55.9) | 51 (50.5) |
| ≥8 months | 1 (1.0) | 0 |
| Smoking: Yes, n (%) | 39 (38.2) | 33 (32.7) |

SARS-CoV-2, severe acute respiratory syndrome coronavirus 2.

^a^Comprised eligible participants who received study intervention with at least one post-vaccination immunogenicity data, and negative anti-SARS-CoV-2 N-protein antibody test result at screening. ^b^≥6 to <7 months is equivalent to ≥180 days to <210 days; ≥7 to <8 months is equivalent to ≥210 days to <240 days; ≥8 months is equivalent to ≥240 days.

**Supplementary Table 2:** Geometric mean fold rise and seroresponse rate for SARS-CoV-2 neutralizing antibodies and anti-spike protein IgG antibodies in the study groups

| **Outcome** | **Tozinameran (n=101)** | | | **S-268019-b (n=101)** | | |
| --- | --- | --- | --- | --- | --- | --- |
| **(95% CI)** | Day 15 | | Day 29 | Day 15 | | Day 29 |
| **Neutralizing antibodies** | |  | | |  | |
| Geometric mean fold rise | 20.77  (18.14, 23.77) | | 16.33  (14.22, 18.76) | 23.34  (20.23, 26.92) | | 22.86  (19.54, 26.74) |
| Seroresponse rate, % | 99.0  (94.6, 100.0) | | 100.0  (96.4, 100.0) | 100.0  (96.4, 100.0) | | 100.0  (96.4, 100.0) |
| **Anti-spike protein IgG antibodies** | | | | |  | |
| Geometric mean fold rise | 38.25  (32.94, 44.42) | | 30.29  (25.92, 35.39) | 35.23  (29.76, 41.70) | | 33.35  (27.93, 39.81) |
| Seroresponse rate, % | 100.0  (96.4, 100.0) | | 100.0  (96.4, 100.0) | 100.0  (96.4, 100.0) | | 100.0  (96.4, 100.0) |

IgG, immunoglobulin G; SARS-CoV-2, severe acute respiratory syndrome coronavirus 2.

The 95% CI were constructed using Student’s *t* distribution for log-transformed titers and the Clopper-Pearson method for proportion data.

**Supplementary Table 3:** Age and neutralizing antibody titer against live wildtype virus in the parent cohorts and in the subgroups

|  | **Tozinameran** | | | | **S-268019-b** | | |
| --- | --- | --- | --- | --- | --- | --- | --- |
|  | **Parent^a^**  **(n=99)** | | **Subgroup^b^**  **(n=24)** | | **Parent^a^**  **(n=100)** | | **Subgroup^b^**  **(n=24)** |
| **Age, years** | |  | |  | |  | |
| Mean | 33.2 | | 32.9 | | 32.7 | | 32.6 |
| Median (range) | 31.0 (21-60) | | 29.0 (21-54) | | 30.0 (21-59) | | 29.5 (22-55) |
| **Neutralizing antibody titer against live wildtype virus on day 29^c^** | | | | | | | |
| Geometric mean | 112.7 | | 113.1 | | 125.5 | | 127.0 |
| Median (range) | 160.0 (20-640) | | 160.0 (20-320) | | 160.0 (20-640) | | 160.0 (20-320) |

^a^The immunogenicity subset excluding participants who tested positive for the COVID-19 polymerase chain reaction (PCR) during the study. ^b^Subgroups were randomly sampled from the immunogenicity subset for each study group excluding participants who tested positive in COVID-19 PCR test during the study. The sampling condition required the immunogenicity subset and the sampled subgroup to be matched for age and day 29 neutralizing antibody titer against live wildtype virus with the Wilcoxon rank sum test (All *P*-values >0.75). ^c^Neutralizing antibody titer was obtained from cytopathic effect-based virus neutralization test.

**Supplementary Table 4:** Incidence of treatment-emergent adverse events by study groups

|  | **Tozinameran (n=103)** | **S-268019-b (n=101)** |
| --- | --- | --- |
| Treatment-emergent AEs | 101 (98.1) | 97 (96.0) |
| Deaths | 0 | 0 |
| Treatment-emergent serious AEs | 0 | 0 |
| Any treatment-emergent AEs of special interest | 0 | 0 |
| Any treatment-related AEs | 101 (98.1) | 97 (96.0) |
| Medically attended treatment-related AEs | 4 (3.9) | 1 (1.0) |
| Nervous system disorders | 43 (41.7) | 26 (25.7) |
| Headache | 43 (41.7) | 25 (24.8) |
| Gastrointestinal disorders | 11 (10.7) | 9 (8.9) |
| Nausea | 5 (4.9) | 5 (5.0) |
| Diarrhea | 6 (5.8) | 4 (4.0) |
| Musculoskeletal and connective tissue disorders | 54 (52.4) | 48 (47.5) |
| Myalgia | 50 (48.5) | 40 (39.6) |
| Arthralgia | 11 (10.7) | 8 (7.9) |
| General disorders and administration site conditions | 89 (86.4) | 79 (78.2) |
| Vaccination site pain | 75 (72.8) | 67 (66.3) |
| Fatigue | 56 (54.4) | 43 (42.6) |
| Pyrexia | 61 (59.2) | 39 (38.6) |
| Vaccination site erythema | 10 (9.7) | 5 (5.0) |
| Chills | 7 (6.8) | 4 (4.0) |
| Investigations | 85 (82.5) | 82 (81.2) |
| Neutrophil percent increased | 80 (77.7) | 77 (76.2) |
| C-reactive protein increased | 46 (44.7) | 33 (32.7) |
| WBC count increased | 11 (10.7) | 9 (8.9) |

AE, adverse event; WBC, white blood cell.

Data are presented as number (%) of participants, where incidence is >1.

A treatment-related AE is defined as an AE considered to be “related” to the study intervention. Participants with multiple treatment-related AEs were counted only once within a system organ class and preferred term.

**Supplementary Table 5:** Incidence of solicited local and systemic treatment-related adverse events over time in the study groups

| **Time (days)^a^** | **Tozinameran (n=103)** | | | | **S-268019-b (n=101)** | | | |
| --- | --- | --- | --- | --- | --- | --- | --- | --- |
|  | **Day 1** | **Day 2** | **Day 3** | **Day 6** | **Day 1** | **Day 2** | **Day 3** | **Day 6** |
| Any systemic solicited TRAEs | 44 (42.7) | 62 (60.2) | 2 (1.9) | 2 (1.9) | 36 (35.6) | 48 (47.5) | 2 (2.0) | - |
| Fatigue | 16 (15.5) | 40 (38.8) | - | - | 24 (23.8) | 19 (18.8) | - | - |
| Fever | 19 (18.4) | 42 (40.8) | - | - | 9 (8.9) | 29 (28.7) | 1 (1.0) | - |
| Myalgia | 20 (19.4) | 30 (29.1) | - | - | 22 (21.8) | 18 (17.8) | - | - |
| Headache | 16 (15.5) | 25 (24.3) | 1 (1.0) | 1 (1.0) | 5 (5.0) | 19 (18.8) | 1 (1.0) | - |
| Arthralgia | 3 (2.9) | 9 (8.7) | - | - | 2 (2.0) | 6 (5.9) | - | - |
| Nausea/vomiting | 2 (1.9) | 2 (1.9) | - | 1 (1.0) | 3 (3.0) | 2 (2.0) | - | - |
| Diarrhea | 2 (1.9) | 2 (1.9) | 1 (1.0) | 1 (1.0) | - | 3 (3.0) | 1 (1.0) | - |
| Chills | 3 (2.9) | 4 (3.9) | - | - | 3 (3.0) | 1 (1.0) | - | - |
| Any local solicited TRAEs (at the injection site) | 42 (40.8) | 37 (35.9) | 1 (1.0) | - | 37 (36.6) | 33 (32.7) | - | - |
| Pain | 42 (40.8) | 34 (33.0) | - | - | 36 (35.6) | 30 (29.7) | - | - |
| Erythema/redness | 5 (4.9) | 4 (3.9) | 1 (1.0) | - | 2 (2.0) | 4 (4.0) | - | - |
| Swelling | - | 1 (1.0) | - | - | - | 1 (1.0) | - | - |

TRAEs, treatment-related adverse events.

Data are presented as number (%) of participants. No participants reported solicited TRAEs on days 4, 5, 7, or 8.

^a^Days after the study intervention dose: (Date of onset) − (Date of study intervention dose) + 1.

If a participant experienced the same AE more than once on different days, then the participant was counted for each day.
